# A feasible and more efficient SARS-Cov-2 vaccine allocation to states and counties in the USA

**DOI:** 10.1101/2021.03.17.21253793

**Authors:** Anthony R. Ives, Claudio Bozzuto

## Abstract

While discussion of vaccine allocation has centered around who should be prioritized (e.g., health care personnel and the elderly), we argue that vaccines should also be allocated to jurisdictions (e.g., counties within the USA) with the greatest immunization thresholds needed for ending the epidemic. At the current rate of vaccine distribution (March 15, 2021), universal herd immunity in the USA could be reached in roughly 4.5 months. However, distributing vaccines according to where the virus spreads more easily (dense counties with high *R*_0_ values), herd immunity would be reached simultaneously in all counties almost two months earlier and would require roughly 40% fewer vaccine doses. Furthermore, under the current distribution strategy densely populated counties would reach herd immunity last, with negative epidemiological and socio-economic consequences. In sum, it would be more fair and efficient to distribute vaccines to jurisdictions that need them most to reach herd immunity.

---

The distribution of vaccines to protect against coronavirus SARS-CoV-2 started in the USA in December, 2020. Because supplies are limited, priorities for the allocation of vaccines have been extensively discussed (*1-2*). The Advisory Committee on Immunization Practices recommended phases for the initial distribution of vaccines (*3*). Highest priority is given to health care personnel who both experience greater risk of contracting COVID-19 and pose greater risk of transmitting the disease to others (phase 1a). These are followed by essential workers and people who are at greater risk of hospitalization and death due to age and/or co-morbidities (phase 1b).

While these recommendations identify who should be given priority for vaccination, they do not explicitly address where the vaccine doses should be distributed. Priorities for the geographic allocation of vaccines need not conflict with priorities based on risks within communities. Instead, geography adds a new dimension to designing fair and effective allocation strategies.

Currently in the USA, vaccines for COVID-19 are being distributed to states in proportion to the number of people living there (*4*). Since this distributes the vaccine evenly among all citizens, it might appear to be fair. However, after protecting the most exposed and vulnerable people, the second important aim of vaccines is to secure herd immunity, which will ultimately allow life to get back to normal. We now know that at the start of the pandemic urban population centers had the highest rates of COVID-19 spread, leading to high estimates of the basic reproduction number (*R*_0_) in counties with high population densities (*5*). Counties with higher *R*_0_ values require higher levels of vaccination to achieve herd immunity (1 − 1/*R*_0_). Therefore, if vaccines were distributed according to *R*_0_, herd immunity throughout the USA would be achieved roughly at the same time in all counties, but also sooner than with the current allocation strategy.

To put numbers on this, we can follow the proportion of the US population living in counties reaching herd immunity through time as vaccines are being distributed (Fig. 1a). Every day each county *i* is proportionally allotted vaccines (*v*_*i*_), either based on its population size (*N*_*i*_),

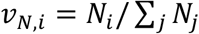

with *j* indexing all counties (*6*), or based on its population size weighted by the proportion of the population needed to be vaccinated to reach herd immunity,

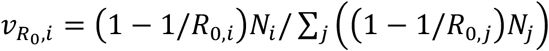

(*5*). For both cases, county *i* reaches herd immunity when a proportion 1 − 1/*R*_0,*i*_ of its population is immune (due to vaccination). To project the time course of counties reaching herd immunity, we assume that (i) the vaccination rate (the first dose of the Pfizer-BioNTech and Moderna vaccines, or the single-shot Johnson & Johnson vaccine) is 1.58 million doses per day, starting from March 15, 2021 (day 0 on the x-axis in Fig. 1) when 71.1 million people had already been vaccinated with at least a single dose (*7*); (ii) vaccines are 90% effective; and (iii) vaccinated individuals cannot spread COVID-19.

**Figure 1.**
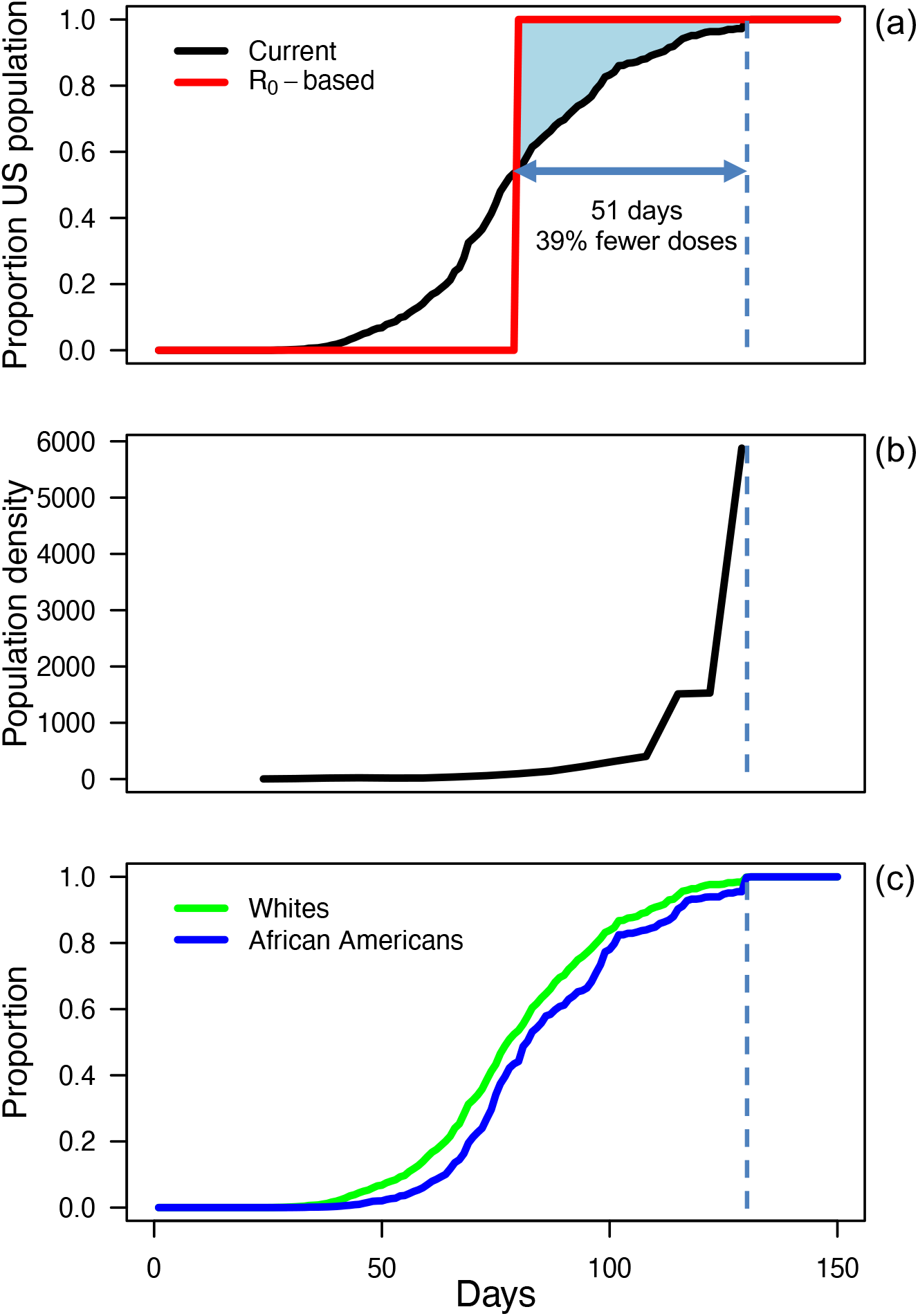
A feasible and efficient allocation of SARS-Cov-2 vaccines according to *R*_0_. Panel **(a)** shows the proportion of the US population living in counties having reached herd immunity, computed as 1 – 1/*R*_0_ at the county level; day 0 on the x-axis is March 15, 2021. The black line assumes the current distribution in which vaccines are sent evenly to counties according to population size, and the red line assumes the distribution depends on estimated county-level *R*_0_ values. Panel **(b)** shows the average population density of counties (individuals per km^2^) that reach herd immunity in a weekly time window. Panel **(c)** gives the proportions of white and African American communities living in counties that have achieved herd immunity.

At the current rate of vaccine distribution, universal herd immunity in the USA could be reached in roughly 130 days with the current population size-based distribution strategy (Fig. 1a). However, the alternative strategy distributing vaccines according to *R*_0_ would mean that herd immunity is reached in all counties at the same time, about 51 days earlier than the last county under the current distribution. Furthermore, 39% fewer vaccine doses would be required to achieve universal herd immunity if vaccines were distributed according to *R*_0_. These results are contingent on the *R*_0_ -based strategy starting mid-March; the overall benefits will expectedly shrink the longer the population size-based strategy is kept.

The exact benefits of distributing vaccines according to *R*_0_ depend on future changes that affect the pandemic. Nonetheless, the savings in time and doses are similarly high under other assumptions. For example, if new virus variants with higher transmissibility become widespread, higher overall vaccination coverage will be needed (*8*). Therefore, we considered the case in which transmission is 1.5 times higher than it was at the start of the pandemic in 2020 (*5*) by setting a new vaccination threshold level to 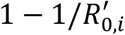, where 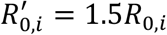. In this case, distributing vaccines according to *R*_0_ would reduce the time to reach herd immunity by 76 days and require 35% fewer vaccine doses.

Under the current distribution strategy, counties with highest density will be the last to reach herd immunity (Fig. 1b), and this has consequences for the current allocation strategy beyond the speed of reaching herd immunity. From an epidemiological perspective, because differences in *R*_0_ values among counties are partly driven by their population density, counties with the highest risk of resurgent and severe outbreaks are last at reaching herd immunity (*5*). From a socio-economic perspective, these high-density counties include many of those that are economic engines in the USA. Finally, high-density counties have relatively large African American populations (*6*), and the current distribution plan leads to a lag of about 4 days behind whites in reaching herd immunity (Fig. 1c).

Despite the current vaccine distribution strategy being well-intended, it would be more fair and efficient to distribute vaccines to states and counties that need them most to reach herd immunity.

## Data Availability

All data used in this study are published and referenced in the main text.

https://doi.org/10.6084/m9.figshare.13322882.v1

http://measureofamerica.org

## Acknowledgments

This work was supported by NASA-AIST-80NSSC20K0282 (A.R.I).

## Data availability

All data used in this study are published and referenced in the main text.

